# The Feasibility of a Smartphone-based Digital Therapeutics for Dysarthria after Stroke (DiDaS): Protocol for a pilot randomized controlled trial

**DOI:** 10.1101/2022.08.16.22278857

**Authors:** Yuyoung Kim, Minjung Kim, Jinwoo Kim, Tae-Jin Song

## Abstract

**Background:** Dysarthria is a motor speech disorder occurring from neurological conditions such as stroke. It leads to serious problems in the intelligibility of patients’ speech, affecting their ability to communicate, quality of life, and emotional well-being. While digital therapeutic approaches for rehabilitation of dysarthria are increasing, clinical evidence of the effectiveness of digital therapeutics has been lacking. A mobile application, D-ST01, has the potential to enhance intensive and repetitive speech rehabilitation due to its high treatment accessibility and its incorporation of gamification, tailored feedback, and interactive functions.

**Methods:** In our trial, 60 stroke patients with dysarthria within 30 days following the occurrence of stroke will be recruited. In a 1:1 ratio, participants will be randomly assigned to either the intervention group (using D-ST01 for 60 minutes/day, five days/week along with usual stroke care) or the control group (usual stroke care only). This will be a single-blind study in which researchers will evaluate outcome measurements while masked to treatment allocation. After four weeks of treatment intervention, we will compare speech and psychological changes between the two groups.

**Conclusions:** Our study will evaluate the feasibility of the speech treatment application D-ST01 for patients with post-stroke dysarthria. In addition, it will collect evidence for investigating the future efficacy of a large-scale randomized controlled trial.

**Trial registration number:** ClinicalTrial.gov #NCT05146765

## Introduction

Stroke is one major cause of worldwide mortality and morbidity [1]. Approximately 40% of stroke patients experience disability [2, 3], and more than 50% of acute stroke victims experience dysarthria [4]. Dysarthria is a neurological motor speech disorder caused by weak, slow, or damaged muscles in the speech-production subsystems [5]. Dysarthria can negatively affect speech intelligibility [5] and communication [6], leading to abnormalities in vocal quality, speed, strength, volume, tone, steadiness, or breath control [5]. Difficulty with communication due to dysarthria after stroke poses a considerable barrier to involvement in social activities and negatively affects the quality of life [7]. It can also lead to emotional difficulties [8], including depression and anxiety.

Intervention for dysarthria involves speech rehabilitation strategies tailored to individual needs and goals [9]. Once emergent medical situations have been settled and the patient is neurologically stabled, intensive speech treatment is prescribed. The evidence suggests that early, rigorous, and repetitive speech therapy can facilitate neurological recovery [10, 11]. However, treatment adherence may be negatively impacted by the patients due to the tediously repeating nature of current speech treatment [12]. Furthermore, since speech therapy involves a substantial amount of time and effort of speech-language pathologist, patients may face restrictions on therapeutic resources [13]. Currently, only about one-third of patients receive sufficient speech therapy. The frequency of therapy also varies from patient to patient [14]. In this context, the use of digital technologies in speech therapy is continuously expanding due to its potential benefits [15].

Digital therapeutics (DTx) is an “evidence-based intervention using high-quality software to prevent, manage, or treat a medical disorder or disease” [16]. There are several advantages to using DTx for self-rehabilitation of dysarthria. DTx can provide more intensive and extended self-administered speech therapy through a smartphone. Also, DTx can overcome the barriers of in-clinic care by providing personalized treatment remotely regardless of time and place. Gamified rehabilitation is an emerging therapeutic methodology that offers good motivation, feedback, and interactivity [17] and can provide repetitive and goal-oriented programs tailored to each patient’s capabilities [18]. Moreover, conversational technologies can facilitate repetitive tasks by providing objective and quantified performance feedback [19]. These features will encourage patients to actively engage in the management of their health by reducing boredom and poor compliance issues [20]. In addition, post-stroke dysarthria patients can have difficulty visiting outpatient clinics due to mobility problems, insufficient family support, and distance between homes and clinics. Therefore, DTx can expand access to treatment [21]. A mobile system incorporating intensive, repetitive, and functional task-oriented speech treatment that can be initiated even during the acute phase will be an alternative to conventional rehabilitation.

However, evidence of the effectiveness of these treatments, particularly DTx, for post-stroke dysarthria is limited due to a lack of sufficiently powered and controlled trials [22]. We hypothesize that stroke patients with dysarthria treated with a speech therapy application called D-ST01 for four weeks will show more improvement in dysarthria symptoms than those who do not receive treatment with the D-ST01 application. Thus, our goal is to develop and test the feasibility of smartphone-based speech therapy, D-ST01, for a stroke patient with dysarthria.

## Methods

### Study design

This is a single-blind, pilot randomized controlled trial with stroke patients with dysarthria allotted to the intervention or control group (Fig 1 and Fig 2). All participants will be recruited among stroke patients from a single stroke center (Ewha Womans University Seoul Hospital) in South Korea. The trial will be conducted in accordance with the Declaration of Helsinki [23]. We will thoroughly explain the purpose and the terms of the study to the participants and receive written informed consent. The study received ethics approval from the Ewha Womans University Seoul Hospital Institutional Review Board on December 21, 2021 (Approval number: SEUMC 2021-12-011). The trial is registered with NCT05146765.

**Fig 1.**
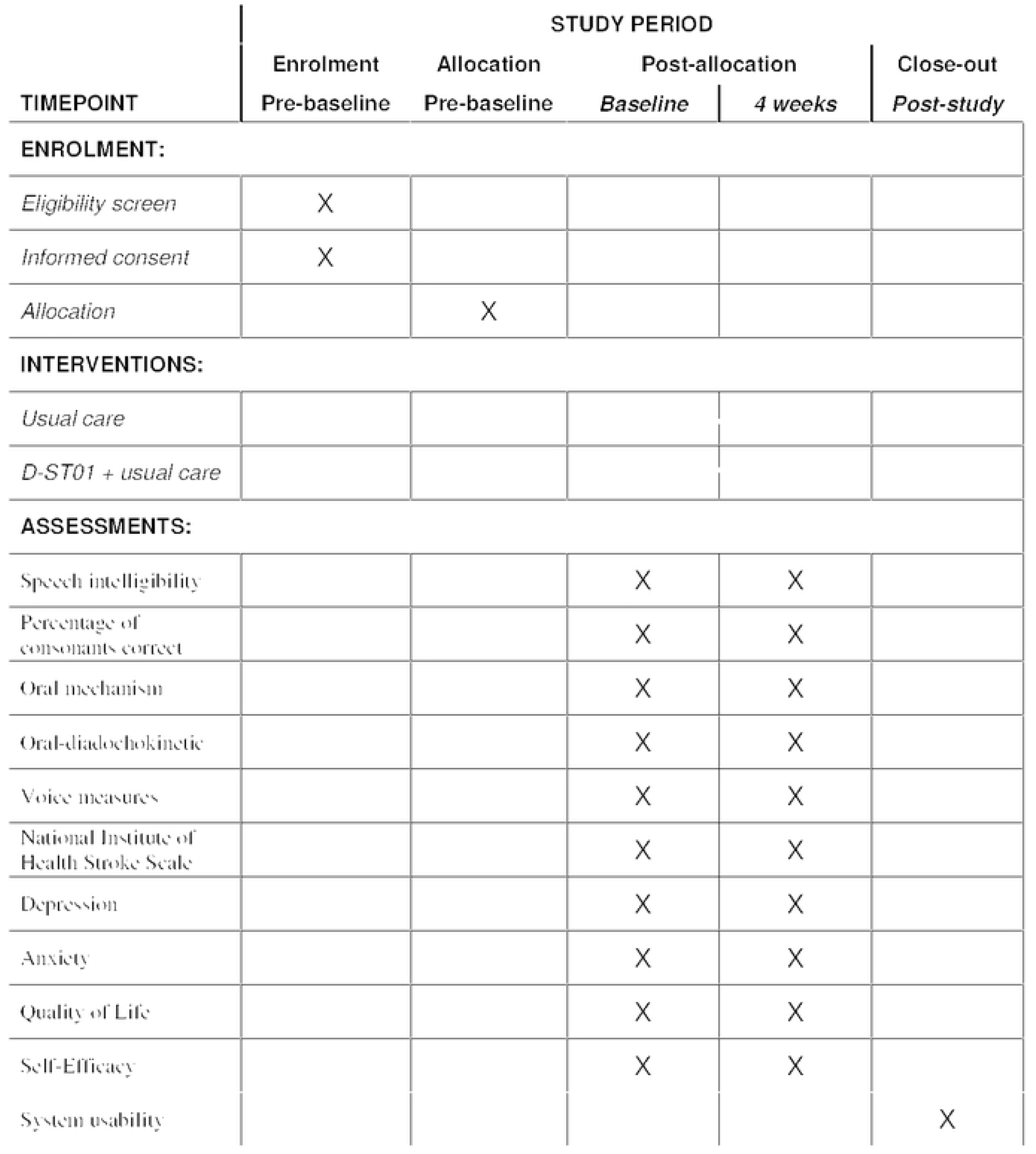
DiDAS pilot trial schedule of enrollment, interventions, and assessments.

**Fig 2.**
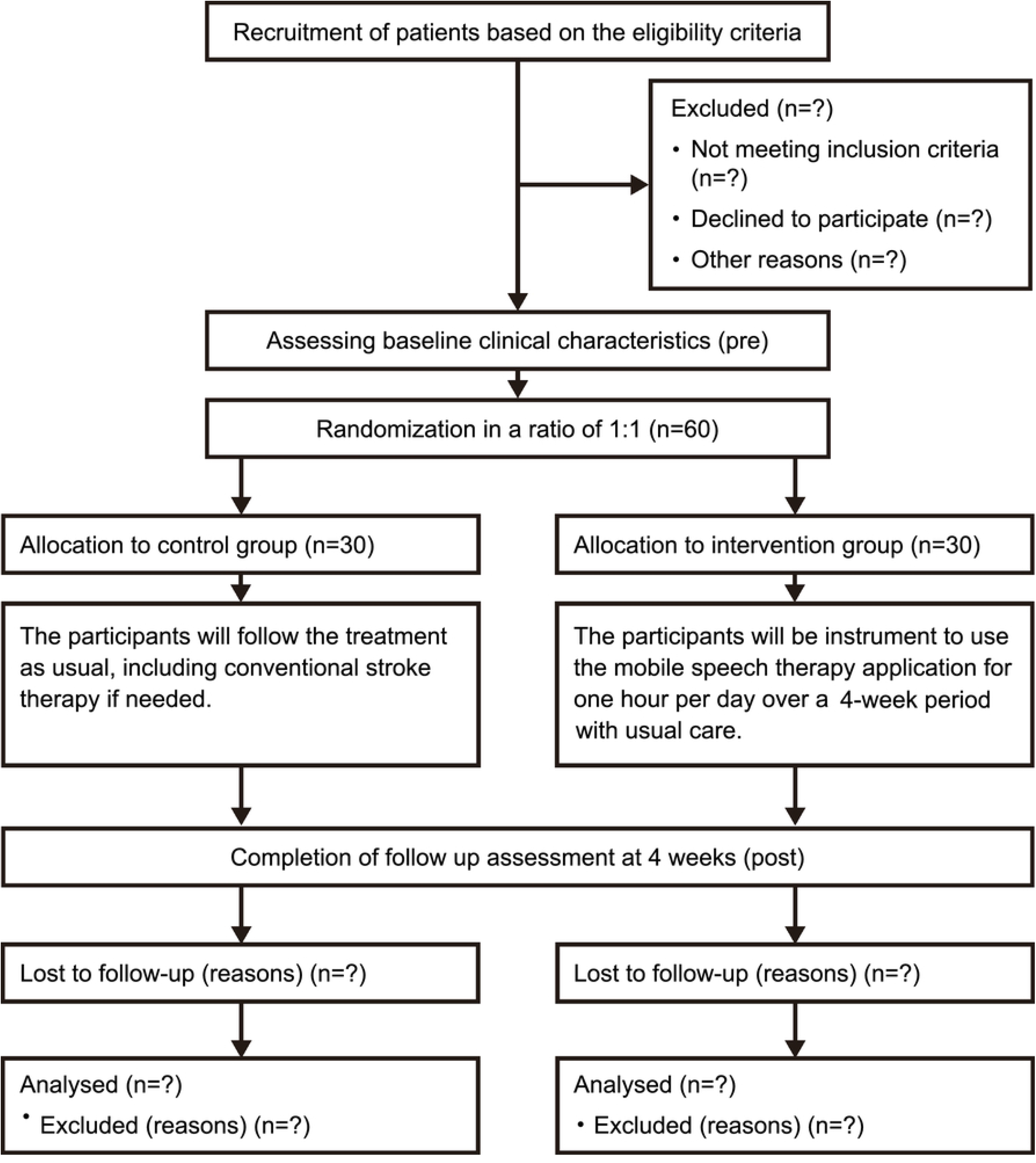
CONSORT diagram representing participant flow in the study.

### Inclusion procedure

The stroke specialist neurologists will screen and enroll participants who meet the eligibility criteria. If a participant meets the eligibility criteria and agrees to participate, a patient information sheet will be delivered to the research coordinator to confirm the patient’s eligibility. The principal investigator will inform the study process to the participants, including the various treatment options available for their symptoms and the risks and benefits of the intervention. Then, the principal investigator will obtain consent from the participant following Research Ethics Committee guidance and Clinical Practice Standards. Table 1 presents this study’s eligibility criteria.

**Table 1.**
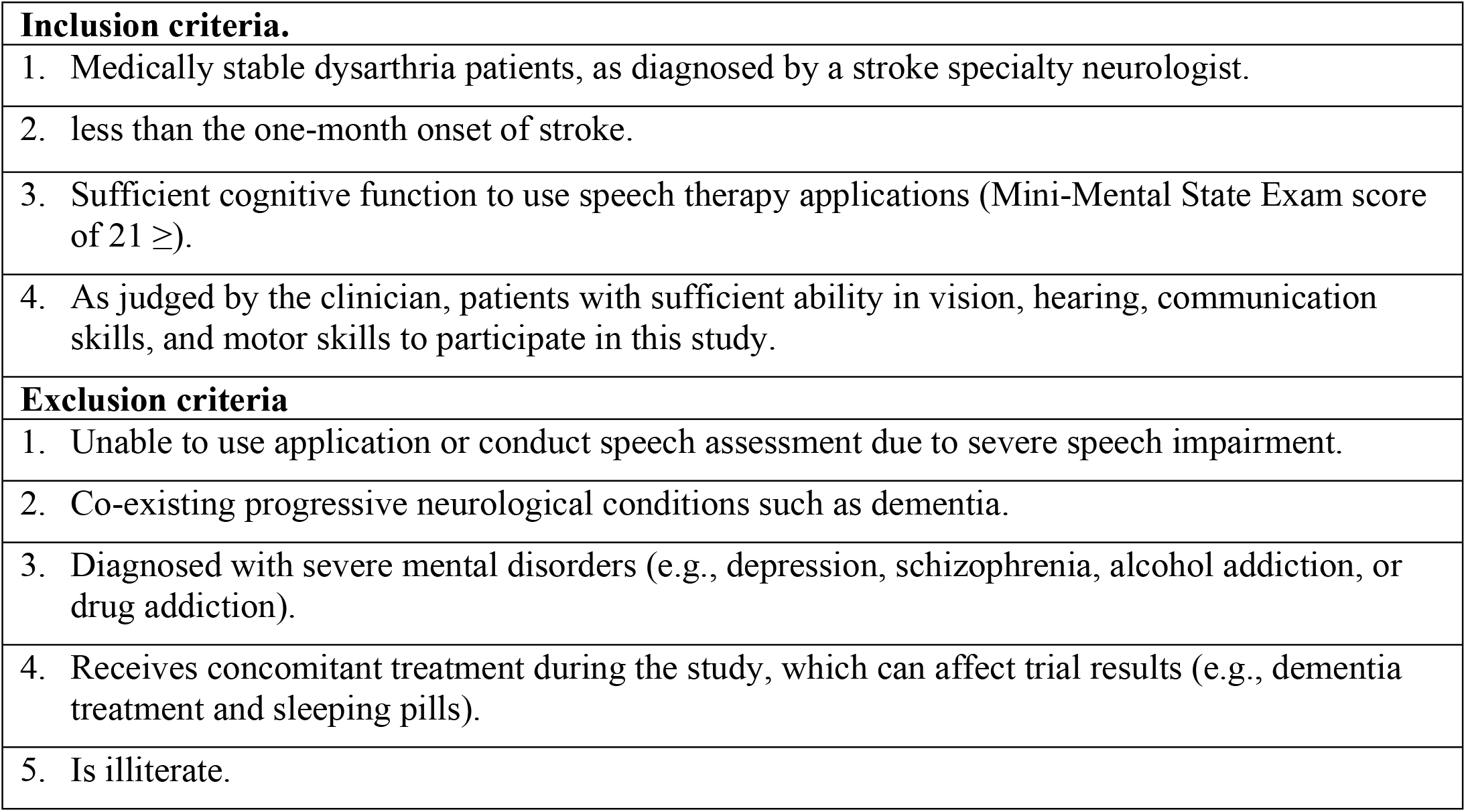
Inclusion and exclusion criteria.

| **Inclusion criteria.** |
| --- |
| 1. Medically stable dysarthria patients, as diagnosed by a stroke specialty neurologist. |
| 2. less than the one-month onset of stroke. |
| 3. Sufficient cognitive function to use speech therapy applications (Mini-Mental State Exam score of $21 \geq$). |
| 4. As judged by the clinician, patients with sufficient ability in vision, hearing, communication skills, and motor skills to participate in this study. |
| **Exclusion criteria** |
| 1. Unable to use application or conduct speech assessment due to severe speech impairment. |
| 2. Co-existing progressive neurological conditions such as dementia. |
| 3. Diagnosed with severe mental disorders (e.g., depression, schizophrenia, alcohol addiction, or drug addiction). |
| 4. Receives concomitant treatment during the study, which can affect trial results (e.g., dementia treatment and sleeping pills). |
| 5. Is illiterate. |

### Randomization

Participants will be randomly allocated with a 1:1 ratio to receive treatment with the speech therapy application along with usual stroke care (the intervention group) or usual stroke care only (the control group). The randomization will be performed by the research officer (not involved in this study) of the clinical trial center of Seoul Hospital, Ewha Womans University. An Excel spreadsheet will be used to generate the random list. The randomization will be created using reasonably sized permuted blocks, and the block size will not be disclosed until the completion of the trial to ensure masking.

### Blinding

Due to the interactive nature of rehabilitation, it is not possible to mask the intervention to patients and research investigators. Therefore, we will blind the separate outcome assessors, who will not be involved in the treatment procedures, to minimize bias. In order to maintain blindness, participants will also be advised not to disclose their assigned group to the outcome assessors.

## Interventions

### Patients allocated to the control group

will receive “usual stroke care” including medical treatment and routine rehabilitation therapy for four weeks. Clinicians and speech-language pathologists will provide usual stroke care based on conventional stroke therapy guidelines [24, 25] and patient needs. This care is expected to be set according to clinical needs as agreed between clinicians and patients and modified according to progress. Participants in this group will only receive usual stroke care and will not use the mobile application, D-ST01.

### Participants in the intervention group

will use the speech therapy application, D-ST01, in addition to receiving usual stroke care (see S3). D-ST01 is a mobile application that delivers speech therapy to patients with post-stroke dysarthria, allowing them to rehabilitate independently. As repetitive and intensive training is crucial in speech therapy, the system features gamification, tailored feedback, and interactive functions designed to motivate patients to persist with constant speech training. Ten different types of speech exercises are provided in the program, with the goal of enhancing speech intelligibility. Participants will perform the speech exercises for one hour per day, five days per week, for four consecutive weeks. They may complete the speech exercises all at once or split them into several sessions. The application provides both written and verbal instructions for each exercise in order to support participants with no or little experience in using a mobile application. The research team will help participants to install the application and will provide a practice session at baseline to enhance participants’ understanding of and engagement with the intervention. Furthermore, participants will be encouraged to reach out to the research team if any issues arise regarding the use of the application.

### Data collection

The primary outcome will be indicated by changes in repeated measured data concerning speech intelligibility between the intervention group and control group from baseline to 4-weeks post-intervention.

Speech intelligibility will be assessed by reading “Gaeul” [26], a text commonly used to evaluate speech disorders in Korea. For future analysis and confirmation, all speech evaluations will be recorded using a high-quality digital recorder (SONY ICD-UX560F). Then, three researchers will listen to the continuous speech of participants and calculate the average by evaluating their intelligibility with a score of 0 (normal) to 6 (unintelligible) [27].

Secondary outcome measures will be evaluated at baseline and after 4 weeks. These will include percentage of consonants correct [28, 29], oral mechanism test [30], oral-diadochokinesis [31], maximum phonation time [32] and phonation evaluation [30]. The experienced speech-language pathologists (with a minimum experience of 6 years), blinded to the participants’ allocation, will be asked to assess various aspects of participants’ speech.

The tertiary outcome will be indicated by changes in a stroke scale and psychological scales. The qualified researchers will evaluate the National Institute of Health Stroke Scale [33]. In addition, we will evaluate patient-reported outcomes concerning depression (Patient Health Questionnaire-9) [34], anxiety (Generalized Anxiety disorder-7) [35], quality of life (Euro Quality of Life) [36], and self-efficacy (Modified Computer Self-Efficacy Scale) [37].

In the last phase of the study, the patients who used the application will complete a system usability scale questionnaire to uncover their experience with the smartphone-based speech therapy [38]. Details of the outcome measures are described in the S2 protocol file.

### Analyses

#### Sample size

The purpose of this study protocol is to investigate the feasibility of digital speech therapy for post-stroke dysarthria, and the results will be used for future power analyses of a larger study. A power calculation was carried out by using G*power 3.0 given the following: 1) power of 80%, 2) a two-sided alpha of 0.05, 3) an estimated effect size of 0.8, and 4) a 1:1 intervention allocation. Thus, each group will need a sample size of 26 participants.

The effect size was calculated by referring to the results of the existing study. Moon and Won studied the effectiveness of smartphone-based speech exercises for 14 acute stroke patients [39]. As a result of calculating the power for the primary outcome with a significance level of 0.05 and 26 participants per group, the oral mechanism was 98%, and oral-diadochokinetic was 98%. Therefore, the use of a major primary outcome, oral mechanism [30], and oral-diadochokinetic [40, 41] for 52 participants are expected to be appropriate in this study. The power analysis is described in detail in S2 protocol file.

A total of 60 participants (30 in each group) will be recruited, considering 15% dropout rate.

#### Statistical methods

All analyses, including a descriptive analysis of demographic and outcome data, will be performed in SPSS (version 25; IBM New York).

An intention-to-treat analysis will be conducted in order to compare outcome measures. First, the Shapiro-Wilk test will be used to examine whether the distribution of continuous variables is normal. Depending on normality, a parametric test (which assumes normal distribution) or non-parametric equivalents (not satisfied with a normality distribution) will be performed. The general characteristics of the two groups will be analyzed with the independent t-test or the Mann-Whitney U test.

Next, paired comparison within the groups (pre- and post-intervention) will be analyzed using the paired t-test or the Wilcoxon signed-rank test. A repeated measures analysis of variance will be performed for each result, taking into account the within-subject factor time (Pre; Post) and the between-subject factor group (Intervention group; Control group). Moreover, a linear mixed model that controls for baseline National Institute of Health Stroke Scale and the location of cerebral infarction as fixed effects will be performed. Statistical significance will be set at p< 0.05. The Last Observation Carried Forward method [42] will be performed to manage the missing data.

#### Monitoring

A data monitoring committee is not needed since there is no risk that mobile-based intervention content will adversely affect participants. Participants may withdraw consent at any time for any reason. Should participants withdraw, they can discontinue the trial without any consequences. Additionally, if a participant requires immediate medical attention, the researchers may withdraw them from the trial.

## Discussion

Our protocol is for a randomized clinical trial of a digital therapeutic provided to stroke patients with dysarthria. Although intensive and repetitive speech therapy is recommended due to its apparent benefits, post-stroke dysarthria treatment remains under-researched [43, 44]. Insufficient methodological rigor, a paucity of studies, and a narrow scope of evaluated interventions limit the available evidence concerning dysarthria treatment [22, 45].

Several studies have adopted traditional speech interventions, including behavioral techniques, breathing, non-speech oro-motor exercises, Lee Silverman Voice Treatment, repetition training of sound, and progressive reading [46]. In a study by Mahler and Ramig [47], reading phrases and Lee Silverman Voice Treatment, which targets high phonatory effort to improve speech skills, were employed in the intervention. Four patients with dysarthria at nine months or more post-stroke received 16 one-hour sessions over four weeks and showed positive responses. Park et al.[48] verified the effect of speech therapy on patients at least six months post-onset of stroke. The intervention consisted of a 60-minute practice, four times per week, for four weeks (16 sessions). The treatment in this study focused on enhancing speech intelligibility with repetitive acoustic-phonetic adjustments.

As in previous studies, our intervention is based on traditional treatments that incorporate motor learning principles (rigorous repetitive training and feedback) [44] and neuroplasticity (repetition, motivation, and reward) [10]. In addition, the treatment duration of one hour, five times per week, for four weeks is consistent with previous behavioral interventions in post-stroke dysarthria [47-50]. However, our protocol differs in enrolling stroke patients from acute to subacute period. The prognosis of speech treatment in the first few months is important since the stroke onset could be critical to reducing recovery time [51]. Nonetheless, the evidence of post-stroke dysarthria in the acute phase is weak [52]. Therefore, the results of our protocol will prove whether the smartphone-based intervention used in this study can help stroke patients with dysarthria to improve their conditions in the acute to subacute period.

As traditional speech therapy continues to fall short of meeting the needs of dysarthric patients, the use of technology in speech rehabilitation has increased, providing new opportunities to overcome the limits of face-to-face treatments. As in all post-stroke rehabilitation, inadequate long-term adherence is a problem in speech therapy [53, 54]. Patients may exercise less frequently than suggested [55], struggle to achieve daily treatment goals [56], or even quit therapy entirely [54]. Digital therapeutics available in home settings are expected to promote high-intensity programs [8], which allow self-management of speech training, facilitate access to care, improve treatment engagement, and lower care costs [16, 57]. Despite the various advantages of technology-based speech therapy, evidence that such therapies are effective in patients with dysarthria is lacking. Our D-ST01 intervention is novel and innovative in that it is composed of several technical features to help with post-stroke dysarthria treatment. D-ST01 adopts gamification elements, responding to users’ speech in real-time and providing points for each task. Since adherence to rehabilitation in stroke patients remains an unmet need, the gamified intervention can make training more enjoyable. As a result, patients’ increased intrinsic motivation can lead to treatment engagement [58, 59]. Technology also enables accurate measurement of several parameters of user interactions and generates valuable feedback to improve the performance of goal-oriented tasks [60]. Participants in this study will receive real-time performance feedback for each exercise (e.g., “Speak louder!”), as well as summary feedback on a daily and weekly basis. The use of this personalized feedback structure is in line with previous findings [44, 61], which indicate that it reinforces positive behaviors and enhances performance. Therefore, the goal of this randomized controlled clinical trial is to identify the feasibility of a smartphone-based post-stroke dysarthria treatment that incorporates gamification, tailored feedback, and interactive functions.

This protocol has a few limitations. First, although this is a pilot trial, the sample size is small. Therefore, generalization may be limited. Second, participants do not receive face-to-face treatment during the intervention. Under these circumstances, patients’ motivation might decrease due to the lack of interpersonal interaction, resulting in a higher-than-expected dropout rate [54, 60]. To address this limitation, we will monitor user data periodically and manage participants through intermittent phone coaching and messaging in order to reduce the number of dropouts. Third, since our D-ST01 app runs only in Korean, the result of the study may not be applicable to stroke patients in other countries. In addition, the study has limited generalizability to the population of patients who are incapable of using or accessing smartphone technology. This issue is particularly prevalent in the elderly population, which is the main target of our study, given that people over 60 account for the majority of stroke cases [61]. However, the findings of this study will form a basis for understanding elderly stroke patients and developing future speech rehabilitation programs for the aging population. Despite these limitations, the findings from this study will inform decisions about effect size estimates and power analysis in future studies.

### Dissemination

Our study protocol will convey key findings and implications to stakeholders related to digital therapeutics for post-stroke dysarthria. The results will be used as evidence for large-scale clinical trials in the future and will be submitted and published in peer-reviewed scientific journals and global conferences.

## Data Availability

Data cannot be shared publicly because of confidentiality. Data are available from the Institutional Data Access or Ethics Committee of Ewha Womans University Seoul Hospital (contact via) for researchers who meet the criteria for access to confidential data.

## Supporting information

**S1 Checklist. SPIRT 2013 checklist.**

(DOCX)

**S2 Protocol.**

(DOCX)

**S3 TIDieR checklist.**

(DOCX)

## Author contributions

**Conceptualization:** Yuyoung Kim, Minjung Kim, Tae-Jin Song

**Methodology:** Yuyoung Kim, Minjung Kim, Tae-Jin Song

**Writing – original draft:** Yuyoung Kim, Minjung Kim

**Writing – review & editing:** Yuyoung Kim, Minjung Kim, Tae-Jin Song

**Supervision:** Yuyoung Kim, Minjung Kim, Tae-Jin Song, Jinwoo Kim

